# COVID-19: Affect recognition through voice analysis during the winter lockdown in Scotland

**DOI:** 10.1101/2021.05.05.21256668

**Authors:** Sofia de la Fuente Garcia, Fasih Haider, Saturnino Luz

## Abstract

The COVID-19 pandemic has led to unprecedented restrictions in people’s lifestyle which have affected their psychological wellbeing. In this context, this paper investigates the use of social signal processing techniques for remote assessment of emotions. It presents a machine learning method for affect recognition applied to recordings taken during the COVID-19 winter lockdown in Scotland (UK). This method is exclusively based on acoustic features extracted from voice recordings collected through home and mobile devices (i.e. phones, tablets), thus providing insight into the feasibility of monitoring people’s psychological wellbeing remotely, automatically and at scale. The proposed model is able to predict affect with a concordance correlation coefficient of 0.4230 (using Random Forest) and 0.3354 (using Decision Trees) for arousal and valence respectively.

**Clinical relevance:** In 2018/2019, 12% and 14% of Scottish adults reported depression and anxiety symptoms. Remote emotion recognition through home devices would support the detection of these difficulties, which are often underdiagnosed and, if untreated, may lead to temporal or chronic disability.

## I. Introduction

The Coronavirus Disease 2019, commonly known as COVID-19, is a viral respiratory syndrome caused by a highly infectious, novel strain of the coronavirus family (SARS-CoV-2) [1]. It emerged at the end of 2019 and rapidly became a pandemic, having an unprecedented impact across the world and leading to the collapse of healthcare systems, with around 150 million cases and over 3 million deaths [2]. In Scotland, the COVID-19 lockdown imposed in March, 2020. The restrictions entailed a ban on all non-essential travel and activities and the advice to work from home and stay at home, in order to slow the spread of the disease and reduce the burden on the NHS. Consequently, everyone’s social and professional lives were seriously disrupted, masks became mandatory, “self-isolation” and “social distancing” made their way into our daily vocabulary and people had to adapt to being confined to their homes. After the summer, cases peaked again and the Scottish government decided to impose a second lockdown just as days were getting darker and colder.

Also in March, the World Health Organisation voiced concerns over the impact of the pandemic on global mental health and published guidelines for self-care, stress, fear management, and dealing with self-isolation [3]. Initial research evidence suggests an increased incidence of anxiety and depression in this period due to persisting stay-at-home orders, financial insecurity, loneliness and overall uncertainty [4].

Early in the pandemic, a Chinese study found a significant drop in life satisfaction and an increased anxiety in 18,000 social media users [5]. Furthermore, 54% of the population in China reported that COVID-19 had a moderate to severe impact on their well-being [6], which was more pronounced in healthcare professionals [7]. Later studies reported similar trends in Europe, America and Oceania. A German study found that 50% of the population suffered from anxiety and spent several hours per day thinking about the pandemic [8]. In Spain, an online study with 3,480 people found increased rates of depression (18.7% participants), anxiety (21.6%) and post-traumatic stress disorder (PTSD; 15.8%), for which loneliness was the strongest risk factor [9]. A Brazilian study also found that 20% of their participants reported severe distress [10]. In the U.S., a survey found increased levels of anxiety and depression as well as increasing financial and health concerns, with especially severe implications for older adults [11]. In New Zealand, a comparable survey found substantial percentages of respondents experienced various degrees of psychological distress. Differently to the U.S. study, this survey found young people to be particularly affected, alongside those who had lost their jobs or had a past history of mental difficulties [12]. These studies show evidence that isolation is a risk-factor for mental health difficulties, especially depression and anxiety.

The study reported here took place in Scotland, where depression and anxiety were already a major health concern before the COVID-19 outbreak. In 2016, the Scottish Burden of Disease Study [13] estimated nearly half a million adults over 16 to be suffering from disability due to depression or anxiety. More recently, in the Scottish Health Survey 2018/2019, 12% and 14% of Scottish adults reported at least two depression and anxiety symptoms, respectively [14].

These difficulties are linked with life satisfaction as they reduce one’s ability to feel joy, take pleasure in our activities and have the energy to fulfil our intentions and live a meaningful life [15]. Isolated people are also less likely to receive help for the difficulties they experience [16]. In this context, our work focuses on emotion recognition through voice analysis, in order to assess the feasibility of mental e-health systems. We present an automatic approach that consists of extracting acoustic features from spontaneous speech data collected during the winter lockdown and training a machine learning model to predict participants’ emotions and energy levels.

## II. Related Work

Most work on machine emotion recognition has been done through image and facial processing [17]. However, emotion recognition through speech analysis is progressively gaining momentum [18], especially so in the context of health research [19]. Speech and language carry information about the speaker, including age and gender [20], as well as physiological, behavioural and emotional information [21]. Speech is ubiquitous and may be collected automatically, unobstrusively and with relatively little infrastructure. This has led to increasing research on technologies for personal health monitoring and diagnostic support tools based on automated processing of speech, in which the field of emotion recognition has been especially active over the past decade. These technologies are supported by machine learning and artificial intelligence, which enable a broad range of analysis and recognition tasks [22].

The premise is that, under different emotions, we produce the same utterance with detectable acoustic differences [23]. Since mental health difficulties often progress with emotional changes, there is an opportunity for signal processing to support the detection of depression, anxiety, apathy, and suicidality [24]. For instance, a recent study on adolescents was able to recognise suicidal ideation thorough an analysis of their voice, using only acoustic features [25]. Another recent study found similar results in their attempt to identify suicidal ideation in war veterans [26].

Emotional processing is challenging, partially because of emotional complexity and a degree of overlap across the emotions that are conventionally designated as primary (sadness, happiness, fear, anger, disgust and surprise) [27]. Categorical recognition of these primary emotions through speech has been a major research focus [28], [29], [30], [31], but a dimensional approach is increasingly popular. This considers primary emotions to be interdependent and operationalises them in two dimensions, namely Valence (V) and Arousal (A) [32], [33], [34], [35]. Briefly, Valence refers to how positive or negative an emotional state is, from unpleasant to pleasant; while Arousal refers to how excited and activated such emotional state feels.

In this work we conceptualise emotionality and affect in these two dimensions (A and V) in order to capture the subtleties that may be overlooked by single-emotion labels. Scores for both of these dimensions were obtained by self-rating through an affective slider [36]. We analysed the voices of 109 individuals and built models to predict their affect scores based on their speech, collected during the winter lockdown. Additionally, we analysed these scores in relation to the Hospital Anxiety and Depression Scale (HADS) [37] in order to establish their association with this symptomatology. To the best of our knowledge, this is the first study utilising speech analysis for recognition of emotional dimensions that are supported with a clinically validated tool (HADS).

## III. Method

### A. Dataset

The PsyVoiD^1^ project investigates the relationship between speech and well-being in the context of the COVID-19. The present subset contains 109 participants who completed the study in Scotland during the winter lockdown. Participants were between 26 and 86 years old 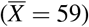 Sixty nine of them are female (63%) and 34 had a past history of depression (31%). All of them completed the HADS questionnaire, which yields anxiety (*M* = 6) and depression scores (*M =* 4). The pleasure and arousal scales range from 100*to*100 (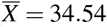 and 30.17, respectively). Table I presents these descriptive statistics.

**TABLE I:** Descriptive statistics on the 109 subjects.

| Variable | Value |
| --- | --- |
| Average age (std) | 59.23 (12.44) |
| Gender (F/M) | 69/41 |
| Past history of depression (Y/N) | 34/71 |
| HADS Anxiety score ($M$) | 6 |
| HADS Depression score ($M$) | 4 |
| Arousal score ($\bar{X}$, std) | 30.17 (52.44) |
| Pleasure score ($\bar{X}$, std) | 34.54 (54.28) |

We report the median ( $M$ ) for HADS due to its skewed distribution (i.e. as this is a non-clinical population, the majority of participants have low scores).

### B. Correlation Analysis

In order to support the clinical validity of the valence and arousal measures, we performed a Pearson’s product-moment correlation between them and the HADS scores. Both were negatively and significantly correlated with HADS (*p* < 0.01), with coefficients of −0.62 and −0.71, respectively. Figure 2 represents this association. Hence, participants with higher scores on the HADS questionnaire reported lower levels in the affective slider. The prompt used to collect the affective scores is shown in Figure 1.

**Fig. 1:**
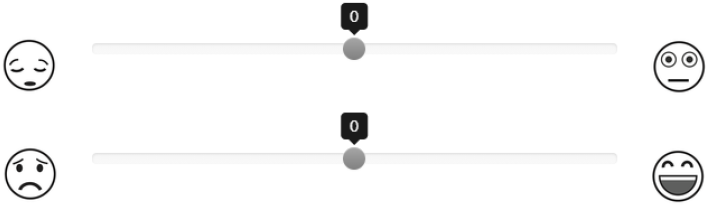
Affective slider: Arousal (top):’How ”energised” do you feel right now?’; and Pleasure (bottom): ’How pleased do your feel at the moment?’.

**Fig. 2:**
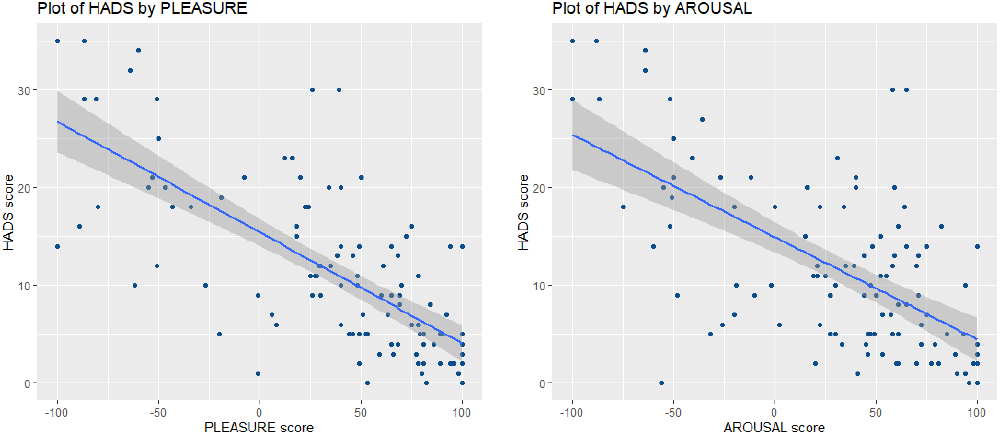
Association between HADS and Affect scores.

### C. Speech processing

The audio files were pre-processed in order to ensure consistency. We implemented spectral substraction for stationary noise removal, audio volume normalisation to control for variable recording conditions, and voice activity detection (VAD) based on signal energy thresholding. The resulting enhanced and segmented recordings were used for acoustic feature extraction and machine learning prediction, and may be made available upon request. There are 3,242 segments from 109 recordings, with an average duration of 72.29 seconds (*sd* = 27.03).

### D. Acoustic feature extraction: eGeMAPS and ADR

The resulting 3,242 acoustic segments were used to extract a comprehensive paralinguistic feature set: *eGeMAPS* [38]. This feature set contains the F0 semitone, loudness, spectral flux, MFCC, jitter, shimmer, F1, F2, F3, alpha ratio, Hammarberg index and slope V0 features, as well as their most common statistical functionals, totalling 88 features per 100ms frame. Features were chosen given their theoretical significance and potential to detect physiological changes in voice production. The *eGeMAPS* set has also proven useful for detection of medical conditions in previous studies [39]. Using the segment level acoustic information extracted, we applied the active data representation method (ADR) to generate a data representation for each audio recording. ADR employs self-organising maps to cluster the original acoustic features and then computes second-order features over these to extract new features. It has been tested previously for large scale time-series data (see [39] for details).

### E. Regression Analysis

We used five types of regression models, namely, linear regression (LR), Decision Trees (DT; where leaf size is optimised through grid search between 1 and 20 and CART algorithm), support vector regression (SVR; with a linear kernel, box constraint *k* optimised though grid search over *k ∈* {10^5^, 20^5^, ….10^6^}, and sequential minimal optimisation solver), Random Forest regression ensembles (RF; where leaf size is optimised through grid search between 1 and 20 with 10 trees in the forest), and Gaussian process regression (GP; with a squared exponential kernel). The regression methods were implemented in MATLAB.

## IV. Results and Discussion

We evaluated a model based on a comprehensive paralinguistic feature set (*eGeMAPS*), a non-linear method for feature extraction (ADR) and five machine learning algorithms for regression. We used the concordance correlation coefficient (CCC) to measure the agreement between the target and predicted scores. CCC is commonly used in emotion recognition and is effectively a non-linear combination of Pearson’s correlation coefficient and the mean square error [40]. The results are summarised in Table II.

**TABLE II:** Leave-one-subject out cross-validation results (CCC)

| Task | LR | DT | SVM | RF | GP |
| --- | --- | --- | --- | --- | --- |
| Arousal | 0.1974 | 0.3841 | 0.3389 | **0.4230** | 0.0215 |
| Valence | 0.1436 | **0.3354** | 0.3131 | 0.3256 | 0.0978 |

Our model was able to predict affect with the best CCC of 0.4230 (RF) and 0.3354 (DT) for A and V, respectively.

While these CCC scores are lower than results reported elsewhere [41] for the SEMAINE dataset (0.680 and 0.506 CCC for A and V, respectively) and the RECOLA dataset (0.692 and 0.423), we note that those datasets consist of recordings taken in carefully controlled conditions and annotated by groups of 6+ annotators, which ensured a level of uniformity unavailable in real-life, self-rated data.

While previous studies typically attempted affect recognition at short (≈ 20*s*) speech segment and/or sentence level [29], [30], [31], this study proposes a system to recognise affect by generating a representation for an entire recording, using segment level information and our ADR method. To our best knowledge, this is the first approach to dimensional affect recognition <u>s</u>ystem using acoustic information for large audio segments (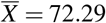 seconds, *sd*= 27.03).

Another relevant limitation of previous studies is the size of the available dataset. There are 10 participants in the EmoDB [29], 4 in the SAVEE dataset [30], 6 in the EMOVO dataset[31], 10 in the vlogger dataset [42]), 23 in the SEMAINE [34] and 46 in the RECOLA [35]. Our study, on the other hand, contains speech of 109 participants and 3,242 segments.

Finally, emotion recognition is often limited by the inherent subjectivity in having emotions labelled by humans who perceive affect from audio, visual and linguistic information [43]. In this study, this intermediate step was removed as the affect is self-reported through the affective slider. In addition, the affective scores were validated by their statistically significant association with the HADS questionnaire (*p* < 0.01, *ρ*_*Arousal*_.= −0.62 and *ρ*_*Valence*_ = −0.71), a long standing tool to screen for depression and anxiety.

## V. CONCLUSIONS

The COVID-19 pandemic has caused great psychological distress among the population. Remote emotion recognition through home devices could support the detection of these difficulties, which are often underdiagnosed and, if untreated, may lead to temporal or chronic disability. Timely identification of these difficulties might enhance the effect of any therapeutic intervention.

This study proposes an affect recognition method with a view to contributing to automatic and remote monitoring of emotional health. Our results evidence the potential to detect emotional and mood difficulties through spontaneous spoken language. In addition, by employing solely on acoustic features, our method is language-independent, and potentially privacy preserving.

Future work will involve analysing linguistic information and its fusion with acoustic information. Other psychological variables available in the dataset will be explored in order to comprehensively assess the potential of our system for mental e-Health.

## Data Availability

Psychological data and enhanced acoustic data may be available upon request (institutional ethical approval for the proposed research will be required).

## Footnotes

1 Ethical approval: June 15, 2020 (REC reference: 20/EM/0146)

